# Prevalence of neutralizing antibody to human coronavirus 229E in Taiwan

**DOI:** 10.1101/2022.03.21.22272725

**Authors:** Hao-Huan Chen, Wei-Fan Chen, Yhu-Chia Hsieh, Chih-Jung Chen

**Author notes:** **Corresponding author**: Dr. Chih-Jung Chen, Department of Pediatrics, Chang Gung Memorial Hospital, No, 5, Fu-Shin St., Kweishan, Taoyuan County 333, Taiwan. Wei-Fan Chen and Hao-Huan Chen contributed equally to this study.

## Abstract

**Background:** Four members in the Coronaviruses family including 229E circulating in the community were known to cause mild respiratory tract infections in humans. The epidemiologic information of the seasonal human coronavirus (HCoV) may help gain insight into the development of the ongoing pandemic of coronavirus disease since 2019 (COVID-19).

**Methods:** Plasma collection containing 1558 samples was obtained in 2010 for an estimate of the prevalence and severity of 2009 pandemic influenza A H1N1 in Taiwan. Of 1558 samples, 200 were randomly selected from those aged < 1 year to > 60 years. The neutralizing antibody titers to HCoV-229E were determined in the serums using live virus ATCC® VR-740^™^ cultivating in the Huh-7 cell line.

**Results:** Seroconversion of HCoV-229E (titer ≥ 1:2) was identified as early as less than 5 years of age. Among 140 subjects aged younger than or equal to 40 years, all of them had uniformly low titers (< 1:10) and the geometric mean titers (GMTs) were not significantly different for those aged 0-5, 6-12, 13-18 and 19-40 years (*P* > 0.1). For 60 subjects greater than 40 years old, a majority (39, 65%) of them had high titers ≥ 1:10 and the GMTs were significantly increased with advanced age (*P* < 0.0001). Age was the most significant factor predicting seropositivity in the multivariate analysis, with an adjusted odds ratio of 1.107 and a 95% adjusted confidence interval of 1.061–1.155 (*P* < 0.0001).

**Conclusion:** HCoV-229E infection occurred as early as younger than 5 years old in Taiwanese and the magnitudes of neutralizing titers against HCoV-229E increased with advanced age beyond 40 years.

## INTRODUCTION

Coronaviruses are positive-sense, single-stranded RNA viruses, causing respiratory or intestinal infections in human beings and/or animals.^1^ They are named for the crown-like appearance under an electron microscope. Before the emergence of SARS-CoV-2 responsible for the ongoing global pandemic of coronavirus disease 2019 (COVID-19), six coronaviruses had been known to cause distinct respiratory syndromes in humans. Four of the human coronaviruses (HCoVs), namely HCoV-229E, HCoV-OC43, HCoV-NL63 and HCoV-HKU1, were seasonal organisms circulating in the community and typically associated with mild and self-limited respiratory tract infections in children.^2^ The other two species including Severe Acute Respiratory Syndrome coronavirus (SARS-CoV-1) and Middle East Respiratory Syndrome coronavirus (MERS-CoV) were of greater virulent potential especially in adult populations and accounted for the epidemics of lethal respiratory diseases in 2002 and 2013, respectively.^3^

It has been suggested that the first seasonal HCoVs infections generally occurred in early childhood and repeated infections with the same virus was frequently identified throughout the whole life.^1,2,4^ However, the infections of seasonal HCoVs appeared to vary substantially across different age groups and in distinct geographic regions. In Taiwan, the HCoV-NL63 was detected in 1.3% of hospitalized children with respiratory tract infections in northern Taiwan during 2004 and 2005 whereas 8.4% of hospitalized patients of all ages with pneumonia in central Taiwan during 2010 and 2011 were associated with this organism.^5,6^ The epidemiologic information of other three seasonal HCoVs were completely lacking in this island. To better understand the epidemiology of seasonal HCoVs, we conducted a seroprevalence study by measuring the neutralizing antibody (NAb) titers against the HCoV-229E among different age groups of Taiwanese population.

## METHODS

### Subjects

The serum samples were selected from the plasma collection containing 1558 samples obtained in 2010, which had been used to estimate the prevalence and severity of 2009 pandemic influenza A H1N1 in the general population in Taiwan.^7^ For this study, the participants were categorized by ages into different groups respectively of age 0 – 5 years, 6 – 12 years, 13 – 18 years, 19 – 60 years and > 60 years. Random samples with indicated numbers as shown in Table 1 were selected from each age group and determined for NAb titers against HCoV-229E virus. The study was approved by the institute review boards in Chang Gung Memorial Hospital (202100266B0). A wavier of inform consent was granted given the retrospective nature of the project and anonymous analysis of the samples and the demographic information.

**Table 1.** Univariate and multivariate analysis of demographics and clinical features in 200 subjects with high and low neutralizing antibody titers against human coronavirus 229E in Taiwan

| Factor | Cases, N (%) |  |  | P value |  |
| --- | --- | --- | --- | --- | --- |
| | Total<br>(n = 200) | High titer<br>($\geq 10$)<br>(n = 39) | Low titer<br>(< 10)<br>(n = 161) | univariate | multivariate |
| Age in years |  |  |  |  |  |
| (Mean ± SD) | 27.0 ± 24.8 | 64.3 ± 9.59 | 17.9 ± 18.0 | < .0001 | < .0001<sup>#</sup> |
| 0-5 | 40 (20) | 0 (0) | 40 (24.8) | < .0001 | ... |
| 6-12 | 40 (20) | 0 (0) | 40 (24.8) |  |  |
| 13-18 | 40 (20) | 0 (0) | 40 (24.8) |  |  |
| 19-40 | 20 (10) | 0 (0) | 20 (12.4) |  |  |
| 41-60 | 20 (10) | 9 (23.1) | 11 (6.83) |  |  |
| > 60 | 40 (20) | 30 (76.9) | 10 (6.21) |  |  |
| Female gender | 123 (61.5) | 21 (53.8) | 102 (63.4) | .2736 | ... |
| Living area |  |  |  | .0005 |  |
| Taipei city | 33 (16.5) | 18 (46.2) | 15 (9.3) |  | Referent |
| New Taipei city | 18 (9) | 2 (5.1) | 16 (9.9) |  | .0455<sup>*</sup> |
| Taoyuan city | 68 (34) | 7 (17.9) | 61 (37.9) |  | .1814 |
| Tainan city | 81 (40.5) | 12 (30.8) | 69 (42.9) |  | .4266 |
| No. of family members<br>(mean ± SD) | 4.59 ± 1.74 | 3.93 ± 1.65 | 4.73 ± 1.74 | .0234 | .8532 |
| Education level |  |  |  | .3381 | ... |
| Non | 38 (19) | 4 (10.3) | 34 (21.1) |  |  |
| Primary | 47 (23.5) | 8 (20.5) | 39 (24.2) |  |  |
| Lower secondary | 31 (15.5) | 9 (23.1) | 22 (13.7) |  |  |
| Upper secondary | 36 (18) | 8 (20.5) | 28 (17.4) |  |  |
| Tertiary | 41 (20.5) | 10 (25.6) | 31 (19.2) |  |  |
| Comorbidity |  |  |  |  |  |
| None | 181 (90.5) | 27 (69.2) | 154 (95.7) | .0001 | .8865 |
| Heart diseases | 8 (4) | 5 (12.8) | 3 (1.9) | .0240 | .9864 |
| Lung diseases | 4 (2) | 2 (5.1) | 2 (1.2) | .4802 | ... |
| Metabolic disorders | 5 (2.5) | 4 (10.3) | 1 (0.6) | .0372 | .9877 |
| Liver or kidney disorders | 3 (1.5) | 2 (5.1) | 1 (0.6) | .1950 | ... |
| Neurologic disorders | 1 (0.5) | 1 (2.6) | 0 (0.0) | .1950 | ... |
| Immune disorders | 1 (0.5) | 0 (0) | 1 (0.62) | 1.0000 | ... |
| pH1N1 |  |  |  |  |  |
| HAI titers<br>(mean ± SD) | 38.3 ± 39.3 | 26.7 ± 24.4 | 41.2 ± 41.7 | .0393 | ... |
| Seroprotection<br>(HAI ≥ 40) | 72 (36) | 5 (12.8) | 67 (41.6) | .0008 | .7127 |
| Vaccination | 101 (50.5) | 16 (41.0) | 85 (52.8) | .1640 | ... |

Abbreviations: SD, standard deviation; HAI, hemagglutination-inhibition assay; pH1N1, pandemic
influenza A H1N1 in 2009
<sup>#</sup>Odds ratio 1.107 95% confidence interval 1.061–1.155
<sup>\*</sup>Odds ratio 10.818 95% confidence interval .787–148.732

### Neutralization assay

The Huh-7 cell line (JCRB0403) was obtained from Japanese Collection of Research Bioresources Cell Bank and grown in low glucose Dulbecco’s modified Eagle medium (LG-DMEM, Gibco) supplemented with 10% heated inactivated fetal bovine serum (FBS, Biological Industries) and 1% penicillin/streptomycin (P/S, Biological Industries) at 37□, 5% CO2. HCoV-229E (ATCC® VR-740^™^) was cultivated in the same medium but with the FBS concentration reduced to 2% at 35□ at 5% CO_2_. The virus was propagated and quantified in Huh-7 cells by measuring the 50% tissue culture infective dose (TCID50). For neutralization test, sera were serially two-fold diluted in the same medium as used for the virus culture, and mixed 1:1 with HCoV-229E (2000 TCID50/mL) in 96-well-plate. The plates then were incubated at 35□ and 5% CO2 for 2h before addition of Huh-7 cells to the mixtures. The result of the virus infection was determined after 5 days of incubation. All sera were previously inactivated at 56□ for 30 min. A titer of ≥ 1:10 was defined as high titer. Comparison of categorical variables between seronegative and seropositive groups was performed with a chi-square test or with the Fisher exact test where appropriate, whereas differences among the numerical variables were analyzed by two-sample t-test. The general linear model (GLM) procedure was used for comparison of geometric mean titers (GMT) between age groups. Multiple logistic regression analysis was applied to explore factors associated with high NAb titers ≥1:10. Statistic significance was defined as a P value of <0.05 in the tests. The statistics were performed using an SAS 9.3 for windows (SAS Institute, Inc., Cary, NC).

## RESULTS

The distribution of NAb titers against HCoV-229E in 200 participants at different ages is shown in Figure 1A. The comparisons of different age groups regarding to the GMTs of NAb is displayed in Figure 1B. Among 140 participants aged younger than or equal to 40 years, all of them had uniformly low titers (< 1:10, Figure 1A) and the GMTs were not significantly different for those aged 0-5, 6-12, 13-18 and 19-40 years (P > 0.1 for all pairwise comparisons by GLM). On the contrary, for 60 participants greater than 40 years old, a majority (39, 65%) of them had high titers ≥ 1:10 and the GMT were significantly increased in participants aged 41-60 years when compared to the younger participants aged 19-40 years (1.57 versus 4.58, *P* < .0001). The GMT level further elevated significantly in the participants aged > 60 years (Figure 1B).

**Figure 1.**
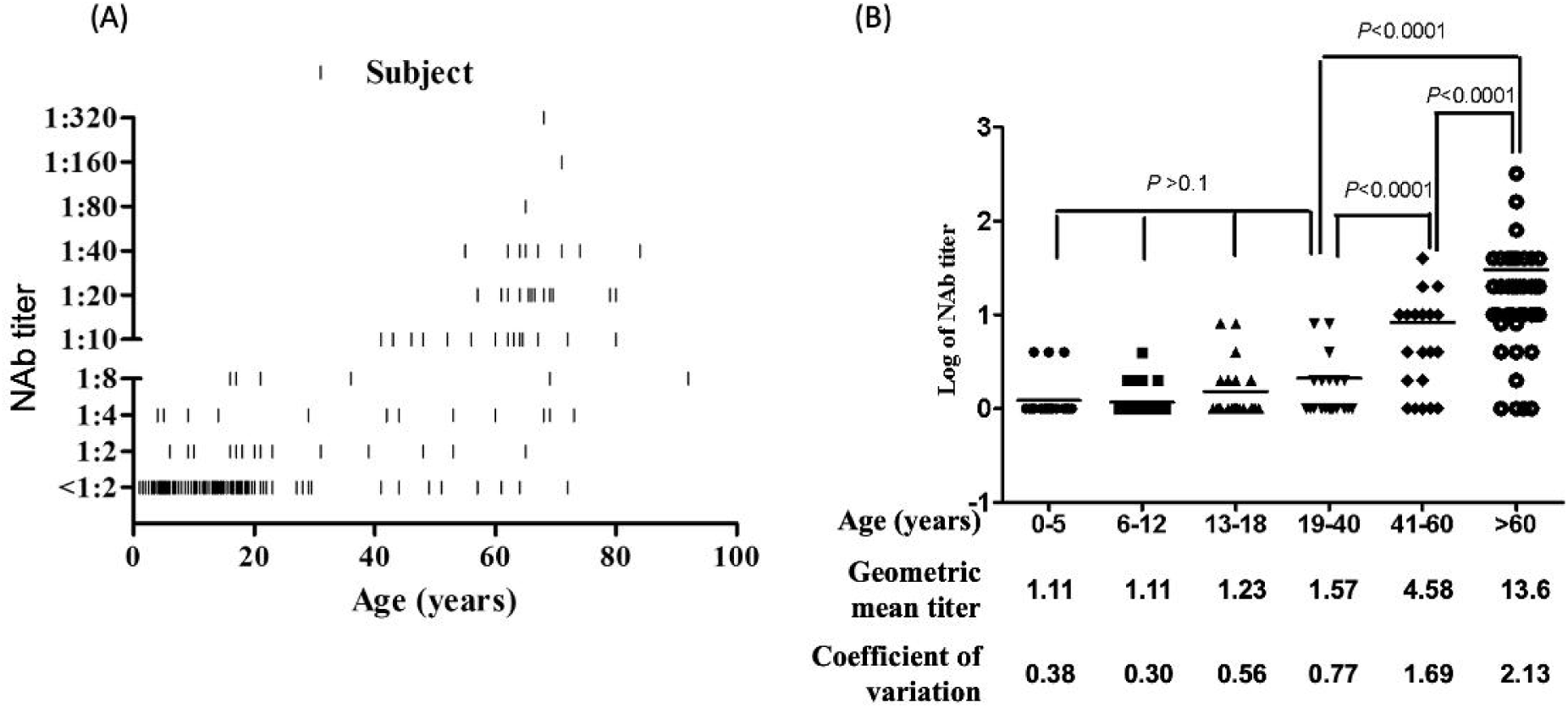
Neutralizing antibody (NAb) titers against human coronavirus 229E. (A) Distributions of NAb titers in 200 Taiwanese at different ages. (B) Comparisons of geometric mean titers of NAb in participants of six age groups. There was no significant difference among four age groups of participants younger than 41 years old (*P* > 0.1 for all pairwise comparisons between the four groups).

To determine the factors associated with elevated NAb titers, we compared a variety of demographic and clinical parameters between the participants by an arbitrarily defined NAb titer cut-off of 1:10. When compared to the participants with low titers (< 1:10), the participants with high titers ≥ 1:10 were at significantly elder age (64.3 ± 9.59 years versus 17.9 ± 18.0 years, *P* < 0.0001, Table 1). In addition to the age factor, the living areas, number of family members, certain co-morbidities including heart diseases and metabolic disorders, and the seropositivity of pandemic influenza H1N1 in 2009 were also significant parameters of participants associated with high NAb titers in the univariate analysis (Table 1). However, a majority of the parameters lost their significance in the multivariate logistic regression analysis. The age remained the most significant factor in the multivariate analysis, with an adjusted odds ratio of 1.107 and a 95% adjusted confidence interval of 1.061–1.155 (*P* < 0.0001, Table 1).

## DISCUSSION

Results from the current study indicated that the age was a significant, if not the only, factor associated with NAb titers against HCoV-229E in Taiwan. It was intriguingly to learn that the NAb titers increased with advanced age, though commenced at young adults, did not reach a significant level until the age of or greater than 40 years old (Figure 1B). The result was inconsistent with the finding of other studies with similar design, which usually disclosed incremental seropositive rates in childhood until a plateau commencing in young adulthood.^1,9^ Ethnic and geographic factors might both be implicated in the discrepant results in the current and others’ studies. Further, the seropositivity was substantially influenced by the sensitivities of different antibody detection methods. Indeed, the detection rates of NAb against HCoVs were usually much lower compared to those determined by the enzyme immunoassay (EIA) methods.^1,8^ No standard method measuring the NAb titers, use of different targets in EIA methods, and no consensus defining seropositivity further complicated the interpretation of the seroprevalence studies. Nevertheless, our study clearly demonstrated the HCoV-229E infection, when defined by a cut-off titer of ≥ 1:2, occurred as early as at 0 – 5 years of age. Before entering into adulthood, at least 17.5% of Taiwanese have been infected with this viral agent and 90% of population had been infected at least once throughout the life. The data indicated that the exposure to HCoV-229E was common in Taiwanese population of all ages irrespective of gender and other demographic characteristics.

The NAb titers against HCoV-229E were at exclusively low levels (< 1:10) in participants younger than 40 years of age. On the contrary, a majority of the cases equal or elder than 40 years of age had high levels of NAb. Of note, the proportions of cases with NAb titer ≥ 1:10 was 45% (9/20) in participants aged 40 – 60 years and increased to 75% (30/40, Table 1) in those elder than 60 years old. The observation might suggest a relatively poor humoral immune response evoked by HCoV-229E infections in young populations. The significant elevation of NAb titer in middle age was likely caused by a more potent or accumulated humoral immunity possibly due to the amnestic response of repeated infections with HCoV-229E. The speculation was supported by a 35-year longitudinal serosurvey investigating the change of antibody titers to HCoVs in 10 individuals in Amsterdam, which demonstrated that the reinfections with the same HCoVs strains indicated by abrupt elevation of antibody titer were common events in adult individuals.^4^

In coincidence with the elevated NAb titers against HCoV-229E in middle age group and elderly, the COVID-19 severity increased significantly in patients beyond 40 years of age. It was suggested that various immune dysregulations were implicated in the severe lung disease of COVID-19. Indeed, an machinery called antibody-dependent enhancement (ADE) was among the potential mechanisms accounting for the enhanced respiratory diseases of COVID-19.^9^ The speculation was supported by the 1-week delayed development of severe respiratory symptoms during the course of COVID-19.^10^ The presence of non-neutralizing antibody during infection has been shown to enhance virus infection, induce uncontrolled cytokine release and Th2 hyperinflammation in the airway with SARS-CoV-1 and MERS-CoV infections. The high antibody titers evoked by the common seasonal HCoVs infections including HCoV-229E might play a contributing role to the greater morbidity and mortality of COVID-19 in patients with advanced ages. Further studies investigating into the interaction between immune profiles of the seasonal HCoVs and SARS-CoV-2 infection might help get insight into the pathogenesis of COVID-19.

In conclusion, the seroconversion of HCoV-229E in Taiwanese occurred early in childhood younger than 5 years of age. Both the positive rates of NAb and magnitude of NAb titers to HCoV-229E increased when the age advanced. The high NAb titer ≥ 1:10 was exclusively identified in middle age group and elderly who were coincidently vulnerable to severe COVID-19 diseases.

## Data Availability

All data produced in the present work are contained in the manuscript

## ACKNOWLEDGMENTS

We thank Dr. Shin-Ru Shih, Director in Research Center for Emerging Viral Infections in Chang Gung University, Taiwan, for providing the HCoV-229E virus.

## TRANSPARENCY DECLARATION

All authors declare no conflict of interest.

